# Smoking, DNA methylation and lung function: a Mendelian randomization analysis to investigate causal relationships

**DOI:** 10.1101/19003335

**Authors:** Emily Jamieson, Roxanna Korologou-Linden, Robyn E. Wootton, Anna L. Guyatt, Thomas Battram, Kimberley Burrows, Tom R. Gaunt, Martin Tobin, Marcus Munafò, George Davey Smith, Kate Tilling, Caroline Relton, Tom G. Richardson, Rebecca C. Richmond

**Affiliations:** MRC Integrative Epidemiology Unit at the University of Bristol, University of Bristol, Oakfield House, Oakfield Grove, Bristol BS8 2BN, UK; Population Health Sciences, Bristol Medical School, University of Bristol, Oakfield House, Oakfield Grove, Bristol BS8 2BN, UK; School of Psychological Science, University of Bristol, 12a Priory Road, Bristol, BS8 1TU, UK; NIHR Bristol Biomedical Research Centre, University Hospitals Bristol NHS Foundation Trust and University of Bristol, Bristol, UK; Department of Health Sciences, University of Leicester, University Road, Leicester, LE1 7RH, UK

## Abstract

Whether smoking-associated DNA methylation has a causal effect on lung function has not been thoroughly evaluated. We investigated the causal effects of 474 smoking-associated CpGs on forced expiratory volume in one second (FEV_1_) in two-sample Mendelian randomization (MR) using methylation quantitative trait loci and genome-wide association data for FEV_1_. We found evidence of a possible causal effect for DNA methylation on FEV_1_ at 18 CpGs (p<1.2×10^−4^). Replication analysis supported a causal effect at three CpGs (cg21201401 (*ZGPAT*), cg19758448 (*PGAP3*) and cg12616487 (*AHNAK*) (p<0.0028). DNA methylation did not clearly mediate the effect of smoking on FEV_1_, although DNA methylation at some sites may influence lung function via effects on smoking. Using multiple-trait colocalization, we found evidence of shared causal variants between lung function, gene expression and DNA methylation. Findings highlight potential therapeutic targets for improving lung function and possibly smoking cessation, although large, tissue-specific datasets are required to confirm these results.

## Introduction

Cigarette smoking is a major risk factor for lung disease, which is often preceded by a rapid decline in lung function ^1^. Studies have shown a strong causal role of smoking in relation to lung function decline, which can be measured by forced expiratory volume in one second (FEV_1_) ^2^. Exploring the mechanistic pathway leading to decreased lung function in smokers may highlight targets for therapeutic intervention.

One possible mechanism which may mediate the association between smoking and decreased lung function is altered DNA methylation patterns. Smoking is associated with substantial changes to methylation levels at many loci across the genome ^3^. For example, hypomethylation at the CpG site cg05575921 in intron 3 of the aryl hydrocarbon receptor repressor (*AHRR*) gene is strongly associated with both current and past smoking behaviour of an individual ^1,4^ and it has recently been suggested to mediate a proportion of the effect of smoking on decreased lung function ^5^. However, it is not clear if this association represents a true causal pathway^6^. Furthermore, DNA methylation at other CpG sites related to lung function may also serve as a potential mediator on the pathway from smoking ^7,8^.

Mendelian randomization (MR) is a method which can be used to assess the causality of a modifiable exposure on an outcome ^9^, using genetic variants robustly associated with the modifiable exposure. As genetic variants are effectively randomized at conception, they are unlikely to be influenced by confounding factors that may otherwise bias associations in observational analysis. In the context of methylation, MR is facilitated by genetic variants found to be strongly associated with DNA methylation, known as mQTLs (methylation Quantitative Trait Loci) ^10^.

Amongst the many extensions of the basic MR principle ^11^ is the two-step method, which aims to assess whether an intermediate factor has a causal role in the mediating pathway between the exposure and the outcome ^12^. A further extension is the two-sample framework, which allows the exposure and outcome data to come from two independent datasets so that the effect of the genetic variant on the exposure and outcome can be estimated separately ^13^. Both approaches are particularly advantageous for epigenetic studies: two-step MR may be used where DNA methylation may serve as an intermediate between a particular risk factor and outcome, and two-sample MR may be used where DNA methylation datasets are unlikely to include the relevant exposure and/or outcome data of interest ^14^. These methods may be used to evaluate the causal role of DNA methylation at a large number of CpG sites, and can be utilised in a mediation framework to determine whether DNA methylation services as a mediator between an exposure and outcome^12,15^. Multiple-trait colocalization can also be used to determine whether variation in DNA methylation levels at putatively causal CpG sites may influence traits via changes in the expression of nearby genes ^16^. Such approaches can be integrated into an analytical pipeline which can be used to highlight and prioritise molecular pathways for further intervention ^14^.

We first aimed to search for a causal effect of methylation at smoking-associated CpG sites on FEV_1_ in UK Biobank using two-sample MR, with replication in the SpiroMeta consortium ^17^. Secondly, we investigated whether DNA methylation mediates the effect of smoking on FEV_1_. Lastly, we assessed whether there is any evidence for shared causal genetic variants between lung function, DNA methylation, and gene expression, using a multiple-trait colocalization framework.

## Results

### Analysis pipeline

A summary of the analysis pipeline used to investigate the causal effect of DNA methylation on lung function is shown in **Figure 1**.

**Figure 1.**
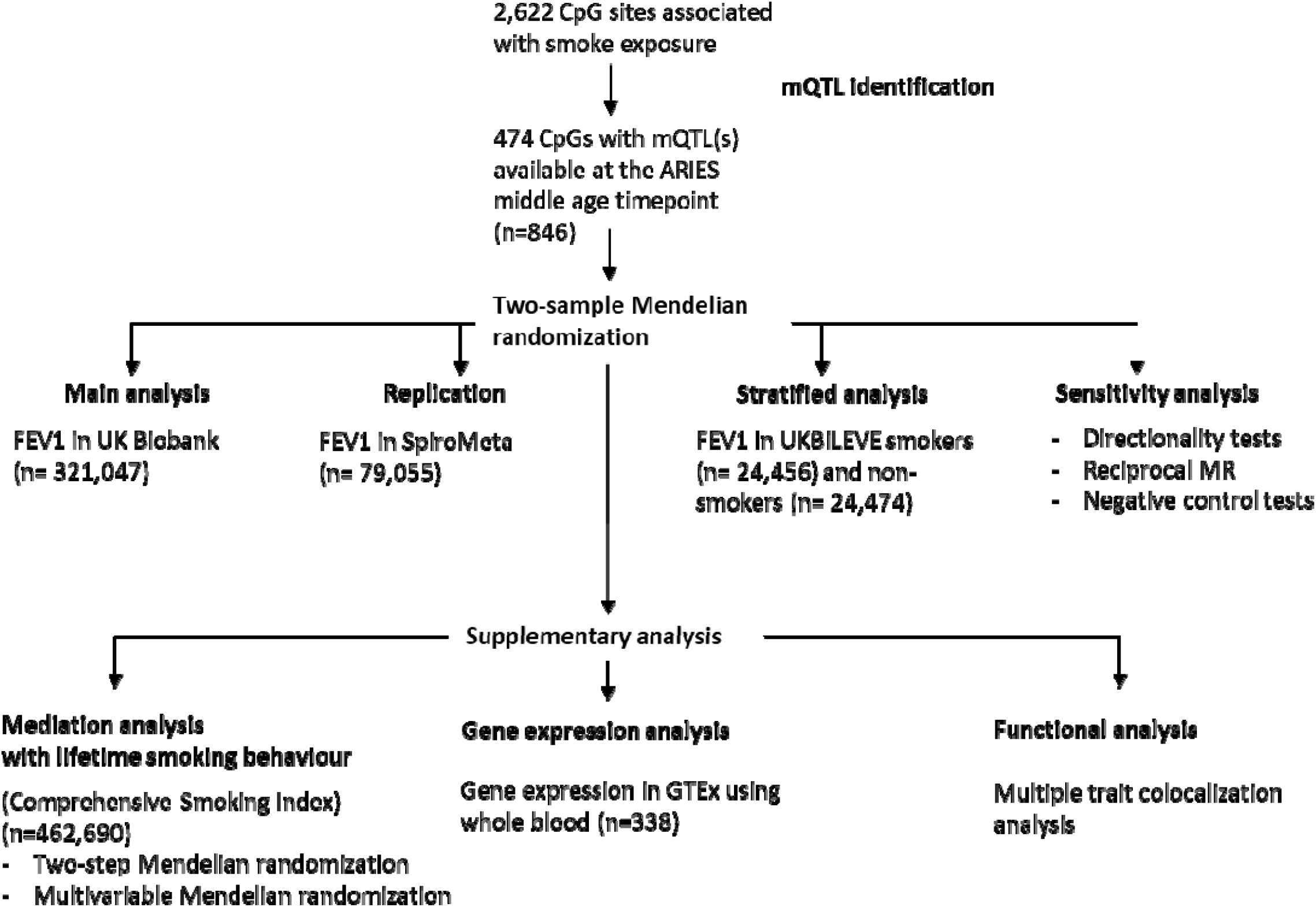
Flowchart of the analysis pipeline, outlining the different analyses performed at each stage of the study. Cohorts and sample sizes used for each analysis are detailed in the flowchart.

### Discovery analysis

We first identified mQTLs which could serve as proxies for 2,622 smoking-related CpG sites identified in a large EWAS meta-analysis conducted by the CHARGE consortium (**Supplementary Table 1**) ^3^. For this, we used a catalog of SNPs associated with CpG sites in the ARIES study (http://mqtldb.org) ^10^ to identify conditionally independent mQTLs (from genome-wide complex trait analysis) from the middle age time point (mean age 47.5 years, n=846) ^10^. There were 474 unique CpG sites associated with smoking (p < 1 × 10^−7^) that we were able to instrument using at least one mQTL (96% in *cis*, 4% in *trans*) (**Supplementary Table 2**). Of these, 415 were present in a GWAS of FEV_1_ conducted in UK Biobank (n=321,047).

To assess the causal effect of DNA methylation at smoking-related CpG sites on lung function, we performed a look-up of the identified mQTLs in the lung function GWAS summary data from UK Biobank and conducted two-sample MR ^13^. We observed 18 CpG-FEV_1_ effect estimates which survived multiple testing correction (Bonferroni p < 1.2 × 10^−4^) (**Table 1, Supplementary Table 3**), with evidence for more causal effects than expected based on chance (**Supplementary Figure 1**).

**Table 1.** Results of two-sample Mendelian randomization analysis of DNA methylation at smoking-related CpG sites on lung function (FEV_1_).

| CpG | chromosome | position | nearest gene | Method | nsnp | b | se | pval |
| --- | --- | --- | --- | --- | --- | --- | --- | --- |
| cg12616487 | 11 | 62379063 | EML3 | Wald ratio | 1 | -0.101 | 0.010 | 3.34E-24 |
| cg09447622 | 6 | 35108605 | TCP11 | Wald ratio | 1 | 0.063 | 0.009 | 4.77E-12 |
| cg21201401 | 20 | 62367884 | ZGPAT | Wald ratio | 1 | 0.076 | 0.013 | 1.54E-09 |
| cg19758448 | 17 | 37828296 | PGAP3 | Wald ratio | 1 | 0.029 | 0.005 | 6.98E-09 |
| cg06382664 | 11 | 73098877 | RELT | Wald ratio | 1 | -0.045 | 0.008 | 1.16E-08 |
| cg24033122 | 16 | 30485383 | ITGAL | Wald ratio | 1 | 0.019 | 0.004 | 2.31E-07 |
| cg09099830 | 16 | 30485485 | ITGAL | Wald ratio | 1 | 0.042 | 0.008 | 2.57E-07 |
| cg21356710 | 2 | 24234017 | MFSD2B | Wald ratio | 1 | 0.030 | 0.006 | 5.19E-07 |
| cg10672416 | 12 | 123718706 | C12orf65 | Wald ratio | 1 | 0.043 | 0.009 | 8.97E-07 |
| cg15059804 | 1 | 33766318 | ZNF362 | Wald ratio | 1 | -0.023 | 0.005 | 2.02E-06 |
| cg10255761 | 3 | 49210029 | KLHDC8B | Wald ratio | 1 | 0.042 | 0.009 | 2.48E-06 |
| cg23771366 | 11 | 86510998 | PRSS23 | Wald ratio | 1 | 0.039 | 0.008 | 2.59E-06 |
| cg09206294 | 15 | 42072687 | MAPKBP1 | Wald ratio | 1 | -0.049 | 0.011 | 2.95E-06 |
| cg15233611 | 12 | 122244660 | SETD1B | Wald ratio | 1 | 0.051 | 0.011 | 3.09E-06 |
| cg19717773 | 7 | 2847554 | GNA12 | Inverse-variance weighted | 2 | -0.032 | 0.007 | 6.94E-06 |
| cg04337534 | 11 | 65816809 | GAL3ST3 | Wald ratio | 1 | 0.052 | 0.012 | 1.16E-05 |
| cg15951188 | 17 | 7832680 | KCNAB3 | Wald ratio | 1 | -0.023 | 0.005 | 3.27E-05 |
| cg11660018 | 11 | 86510915 | PRSS23 | Wald ratio | 1 | 0.036 | 0.009 | 5.54E-05 |

Two-sample Mendelian randomization analysis using SNP-methylation estimates from ARIES (sample 1, supplementary table 2) and SNP-FEV<sub>1</sub> estimates from UK Biobank (sample 2). The effect size (b), standard error (se) and p value (pval) for each CpG reaching significance after Bonferroni is reported, along with the chromosome and position of the CpG, the nearest gene, and the Mendelian randomization method used to analyse the effect on lung function.

Given previous findings of a mediating role of *AHRR* (cg05575921) methylation in the relationship between smoking and lung function ^18,19^, we specifically tested the causal effect of methylation at cg05575921 on FEV_1_ in a MR framework. As there were no mQTLs found to be robustly associated with this CpG site in the middle age time point of ARIES, we identified two mQTLs from the ARIES childhood timepoint which we took forward for MR analysis (**Supplementary Table 4**). This revealed no strong evidence for a causal effect of *AHRR* (cg05575921) methylation on FEV_1_ (**Supplementary Table 5**).

### Replication analysis

We attempted to replicate effect estimates for the top 18 CpG sites identified in UK Biobank using data from the SpiroMeta meta-analysis GWAS of FEV_1_ (n=79,055) (**Figure 2**). Three CpGs replicated beyond a stringent Bonferroni threshold (cg21201401 (*ZGPAT*), cg19758448 (*PGAP3*) and cg12616487 (*AHNAK*)) (p<0.0028) (**Supplementary Table 6**) and there was consistency in the direction of effect at 15 of the CpG sites (83%).

**Figure 2.**
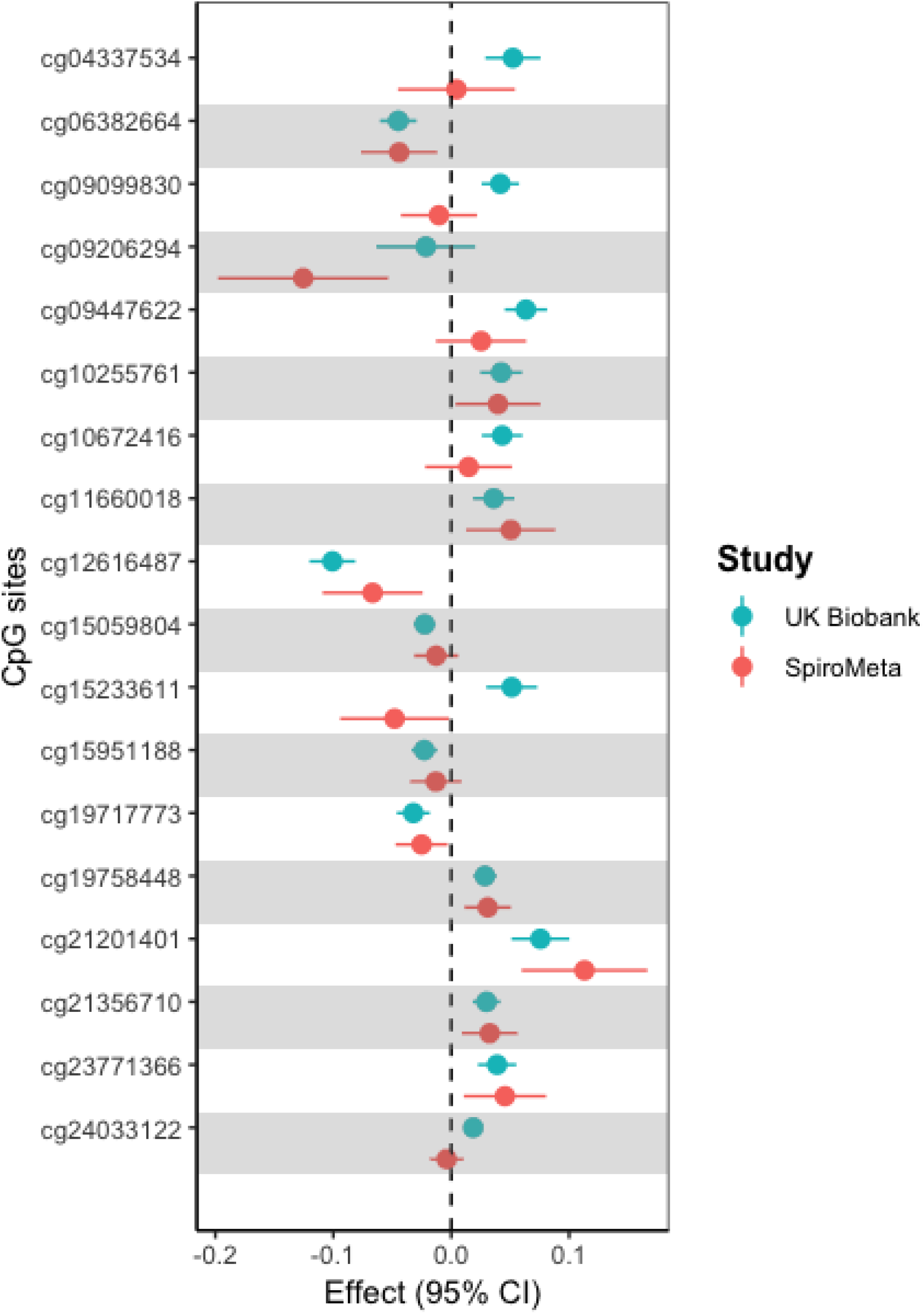
Results of MR analysis of the effect of smoking-associated DNA methylation on lung function (FEV_1_) in UK Biobank (discovery) and SpiroMeta (replication). Effect sizes and 95% confidence intervals (CI) of the eighteen significant CpG sites from the discovery analysis are shown in blue, and the effect sizes and CI of the same CpG sites in the replication analysis in SpiroMeta are shown in red

### Stratified analysis

The sample used in the discovery analysis included current, former, and never smokers in UK Biobank, and so we performed a stratified analysis using never- and heavy-smoking subsets of UK Biobank in the UKBiLEVE dataset. This stratified analysis had less statistical power than the discovery analysis due to a tenfold drop in sample size (smokers, N = 24,457; non-smokers, N = 24,474). The results from the stratified analysis are compared to the discovery analysis in **Supplementary Figure 2**, and **Supplementary Table 7** shows the results for the top CpGs. Effect estimates were generally similar between the mixed, smoking-only and non-smoking only groups. For some sites (cg09099830, cg09206294, cg24033122 and cg10672416) an effect was present in smokers but not non-smokers, while at others (cg10255761, cg21201401 and cg04337534) there was a larger effect in non-smokers compared with smokers.

### Sensitivity analysis

#### DNA methylation and lung function: direction of causality

We performed directionality tests using the MR Steiger method ^20^ to provide evidence that the causal pathway was in the direction from DNA methylation to FEV_1_, rather than vice versa. This was suggested to be the case for all CpG sites in the main analysis (**Supplementary Table 8**), as the mQTLs explained substantially more variation in DNA methylation (between 3.3% for cg09447622 and 31.3% for cg24033122) than in FEV_1_ (r^2^ < 0.04%). When testing the impact of FEV_1_ on DNA methylation, using 175 out of 221 SNPs identified in the UK Biobank GWAS ^17^ as genetic instruments, we found little evidence to suggest that lung function had a causal effect on DNA methylation levels at any of the 18 CpG sites (**Supplementary Table 9**).

#### Smoking behaviour and DNA methylation: direction of causality

We also evaluated the direction of causality between lifetime smoking behaviour and DNA methylation at the identified CpG sites, using 119 out of 126 SNPs identified from a GWAS of a comprehensive smoking index metric ^21^ as genetic instruments. There was limited evidence that lifetime smoking behaviour had a causal effect on DNA methylation at the 18 CpG sites of interest, and the effect estimate from MR analysis was consistent in terms of direction of methylation with the original smoking EWAS at only 12 of the 18 CpG sites (**Supplementary Table 10**), which is in contrast to the majority of the smoking-related CpG sites where the MR estimates were more in line with those from the smoking EWAS (**Supplementary Figure 3**).

Conversely, there was evidence for a causal effect of DNA methylation on lifetime smoking at several of the CpG sites when we performed the reciprocal MR analysis (**Supplementary Table 11**). We also performed directionality tests using the MR Steiger method which provided evidence that the causal pathway was in the direction from DNA methylation to smoking, rather than vice versa (**Supplementary Table 12**).

#### Negative control

Given differences in sample sizes between the DNA methylation and lifetime smoking datasets which may bias the directionality tests ^20^, we also carried out further analysis using mQTL data from the childhood time point (mean age 7.5 years, n=885) as a negative control. We showed that the mQTLs were strongly associated with DNA methylation in the ARIES childhood time point (i.e. in non-smoking individuals) to rule out the possibility that the mQTLs were having their primary effect via smoking (**Supplementary Table 13**).

### Supplementary analyses

#### Mediation analysis

Given the limited evidence to suggest that smoking has a causal effect on DNA methylation at the 18 CpG sites of interest, we conducted mediation analysis to investigate the mediating pathway from DNA methylation to lung function via lifetime smoking behaviour at the 7 CpG sites where there was evidence for a causal effect of DNA methylation on lung function as well as smoking behaviour beyond a Bonferroni threshold (p<0.0028) (**Supplementary Table 11**).

This was done first by performing two-step MR analysis^12^ (**Figure 3**) and using the “product of coefficients” method ^22^ to estimate the indirect effect of DNA methylation on lung function via lifetime smoking. For all 7 CpG sites there was evidence of an indirect effect of DNA methylation on lung function via lifetime smoking (p≤0.006), mediating between 7.85 and 19.33% of the total effect (**Supplementary Table 14**). This indirect effect was replicated for 5 CpG sites when using FEV_1_ GWAS summary data from SpiroMeta (p≤0.008) (**Supplementary Table 15**).

**Figure 3.**
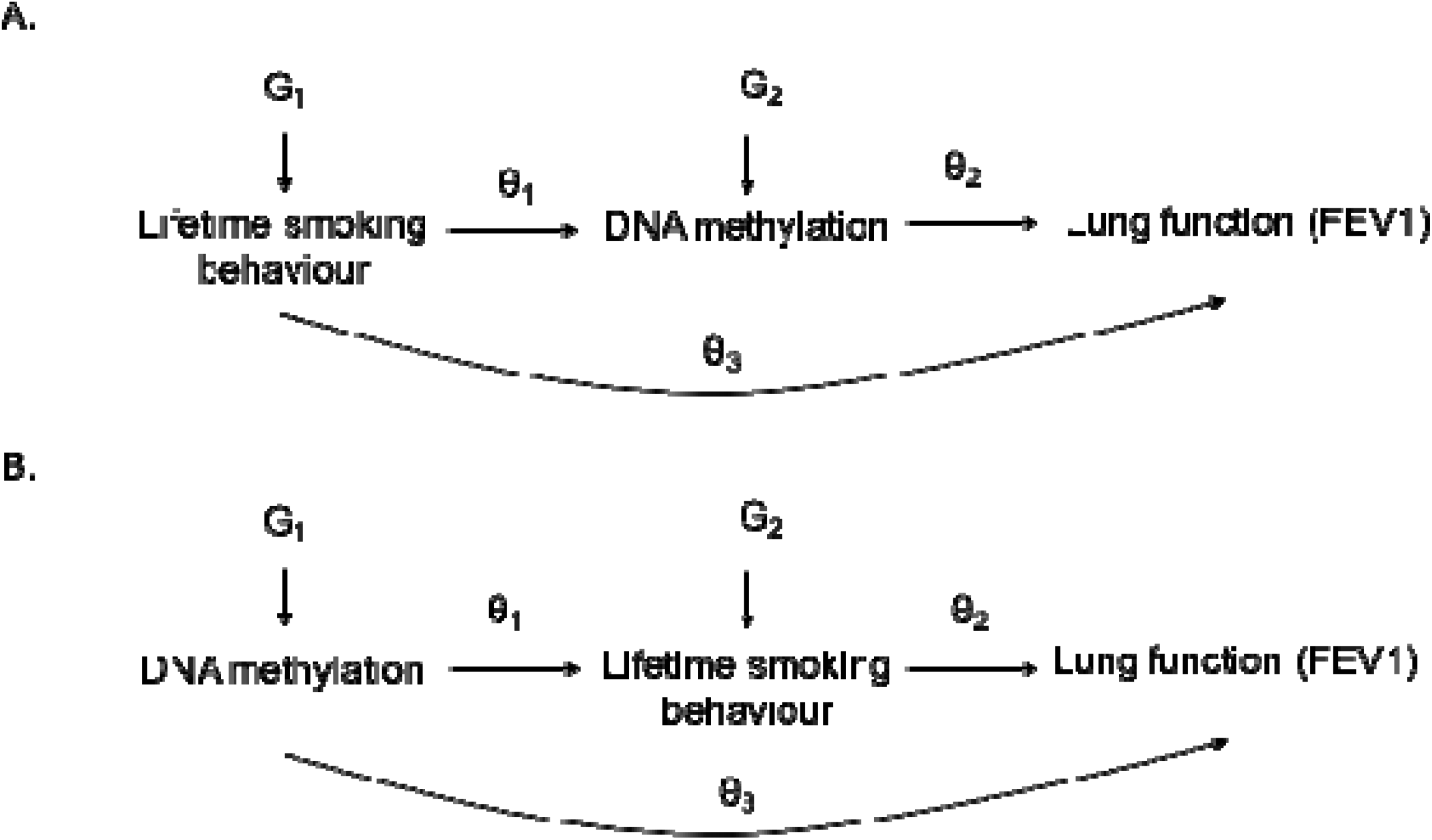
Outline of the steps of the mediation analysis. A. Assessing the mediating role of DNA methylation in the effect of smoking behaviour on lung function (FEV1). B. Assessing the mediating role of smoking behaviour in the effect of DNA methylation on lung function (FEV1). θ_1_ = Step 1; θ_2_ = Step 2; Indirect effect = θ_1_ x θ_2_; Direct effect = θ_3_; Total causal effect = θ_3_ + θ_1_ x θ_2_

We also estimated the direct effect of methylation on lung function using a multivariable MR (MVMR) approach ^23,24^, and used the “difference of coefficients” method ^22^ to determine the indirect effect of DNA methylation on lung function via lifetime smoking (**Supplementary Table 16 and 17**). Although independent instrument strength for lifetime smoking and the 18 CpG sites was deemed to be strong (Q-statistics ≥ 64.5), the indirect effect was estimated with lower precision than the two-step MR analysis. In addition, there was some evidence for heterogeneity in the causal effect estimates from the MVMR which could indicate the presence of invalid instruments (e.g. due to horizontal pleiotropy) ^24^ (**Supplementary Table 17**). Nonetheless, there was supportive evidence for an indirect effect of methylation at two sites (cg10255761 and cg15951188) on FEV_1_ via smoking in MVMR (**Supplementary Table 16 and 17**).

#### Multiple-trait colocalization analysis

For those CpG sites where there was evidence of a causal effect on lung function, we applied a genetic colocalization approach to determine whether the variant responsible for influencing methylation at each CpG site was the same variant influencing changes in lung function. Furthermore, it is likely that any true association between a CpG site and lung function is mediated by changes to the expression of nearby genes. To assess this, we applied multiple-trait colocalization (‘moloc’) ^16^ to investigate whether the variant responsible for influencing methylation at each CpG site was the same variant influencing changes to both nearby gene expression and lung function ^25,26^.

There was strong evidence (based on PPA ≥ 80%) at 5 CpG sites that variation in DNA methylation, gene expression and FEV_1_ were all attributed to the same underlying genetic variant. This included associations at cg21201401 (with *ZGPAT* expression (PPA=84.2%)) and cg12616487 (with *AHNAK* expression (PPA=88.9%), which were two of the CpGs where effects on FEV_1_ most strongly replicated in SpiroMeta. This suggests that the relationship between DNA methylation at these smoking associated CpG sites and lung function may also involve the transcription of nearby genes, which is a mechanism of effect consistent with causality. There was also strong evidence of colocalization at a further 4 CpG sites, although only between DNA methylation and FEV_1_ (but not nearby gene expression). Colocalization results are shown in **Supplementary Table 18**.

## Discussion

We investigated CpG sites previously associated with smoking for their potential causal impact on lung function, using a two-step MR framework. A discovery MR analysis using mQTLs associated with 474 smoking-associated CpGs identified 18 CpGs with a possible causal effect on lung function in UK Biobank. Replication in SpiroMeta provided supportive evidence for a causal effect of methylation on FEV_1_ at three CpG sites and there was consistency in the direction of effect at 83% of the CpG sites. A further analysis stratified by smoking status using the UK BiLEVE dataset highlighted heterogeneity in effects among heavy smokers compared with non-smokers at some of the sites.

We found little evidence to suggest that lung function in turn influenced levels of DNA methylation at the 18 CpG sites. Interestingly, MR analyses provided limited evidence that smoking had a causal effect on DNA methylation at these smoking-related sites. Instead, we observed that at several of the CpG sites DNA methylation had a causal effect on smoking. We therefore conducted mediation analysis using both two-step and multivariable MR to estimate the extent to which smoking mediates the association between DNA methylation and lung function at these sites. In two-step MR, evidence of mediation was found for 7 CpG sites when using FEV_1_ GWAS summary data from UK Biobank and for 5 CpG sites using SpiroMeta. Indirect effects were estimated with less precision in the MVMR approach.

While we were unable to robustly demonstrate that these effects occur along a common causal pathway and the effects observed could be due to horizontal pleiotropy, we performed colocalization to distinguish causal pathways from linkage disequilibrium between causal variants for DNA methylation and lung function. We also integrated evidence from gene expression to provide further evidence for causality.

### Comparison with other studies

We searched both the EWAS Catalog and the EWAS Atlas ^27^ to assess whether any of the 18 CpGs where DNA methylation was identified as having a possible causal effect on lung function in MR analysis have been previously identified in other epigenome-wide association studies of lung function or chronic obstructive pulmonary disease (COPD). The CpG sites cg15059804 (*ZNF362*) and cg11660018 (*PRSS23*) were found to be associated with asthma in an EWAS conducted in lung cells^28^; cg11660018 was also suggested to have a causal effect on lung function, along with cg23771366 (*PRSS23*), in another EWAS conducted in blood which was followed up by a two-sample MR analysis^7^. The direction of causal effect for these two CpGs in this MR analysis was consistent with our results.

One CpG site, cg21201401 (*ZGPAT*), was found to be inversely associated with COPD below FDR correction (beta = -0.091, p = 0.0003) in an EWAS conducted in lung tissue (114 subjects with COPD, 46 controls who were all former smokers) ^29^. This effect is consistent with our observation of a causal effect on increased FEV_1_, which could feasibly provide a protective effect against the development of COPD.

As mentioned in the Introduction, one previous study performed mediation analysis which indicated that hypomethylation at cg05575921 (*AHRR*) might mediate the association between smoking and lung function ^7^. However, we found in MR analysis that there was no strong evidence for a causal effect of *AHRR* methylation on FEV_1_, indicating that it is unlikely to be mediating the effect of smoking on lung function. Similar conflicting findings have been observed between conventional mediation approaches and MR analysis to determine epigenetic mediation, in the context of smoking and lung cancer ^30,31^ and prenatal famine and later life metabolic profile ^32,33^. Traditional mediation approaches are more susceptible to measurement error and potential reverse causation than MR^34^, meaning the proportion of the mediated effect reported by these studies is likely to be overestimated. However, several limitations of MR analysis have also been raised previously which may explain discrepancies in these results, including tissue-specificity, pleiotropy and low power ^35^. These limitations are discussed in turn below.

### Limitations

#### Sample considerations

A possible explanation for why this MR analysis did not detect a causal effect of smoking on DNA methylation is low power due to the small sample size for the DNA methylation sample (n=846). Three of the eighteen sites identified in our analysis as having a causal effect on lung function were also previously implicated in an EWAS of maternal smoking in pregnancy ^36^, although the direction of effect was not always consistent with our results. These were: cg12616487 (*EML3*), cg23771366 (*PRSS23*), and cg21201401 (*ZGPAT*). As DNA methylation is unlikely to directly influence maternal smoking in this instance, this indicates that smoke exposure (own smoking or in-utero) may have a causal effect on DNA methylation which was not detected in our MR analysis.

Furthermore, while both the GWAS for lifetime smoking and lung function were conducted in samples which include both males and females, the mQTL effects used in the main analysis were obtained in females only in ARIES. Nonetheless, we have shown consistency in the mQTL effects in a mixed sample of males and females from the ARIES childhood time point.

An additional sample consideration relates to the use of both UK Biobank and the UKBiLEVE subset, both of which represent selected groups of the population which could bias effect estimates in the MR analysis ^37^. Nonetheless, we have also performed independent replication using data from 22 studies in the SpiroMeta consortium which provided confirmatory causal estimates at the majority of the identified CpG sites.

#### Horizontal pleiotropy

As mentioned, another limitation of the MR approach to assess the causal effect of DNA methylation is the assumption of no horizontal pleiotropy. While various sensitivity analyses exist for investigating horizontal pleiotropy in MR analysis ^38^, these approaches rely upon the existence of multiple genetic instruments for each exposure and therefore the application of these approaches is restricted in the setting of evaluating methylation changes, where few independent mQTLs exist for individual CpG sites. However, we performed a colocalization analysis on our top hits to investigate the relationship between methylation of these sites, expression of nearby genes, and variation in lung function. If all three of these traits were to share a common causal variant, it would suggest that associations are more likely be due to an underlying causal relationship as opposed to genetic confounding (i.e. high linkage disequilibrium existing between an mQTL and variant which influences lung function).

Our colocalization analysis revealed that genetic variation associated with DNA methylation colocalizes with both lung function variation and gene expression at serval sites. For example, methylation at cg21201401 was shown to colocalize with *ZGPAT* expression and lung function, and methylation at cg12616487 was shown to colocalize with *AHNAK* expression and lung function. The *AHNAK* gene is responsible for a neuroblast differentiation-associated protein, which has previously been reported to confer risk of chronic obstructive pulmonary disease (COPD) due to missense variants in its coding region ^39^. This makes *AHANK* a strong candidate responsible for the association with lung function risk at this locus.

Furthermore, MR associations at both cg21201401 and cg12616487 were replicated using data from the SpiroMeta consortium. This further supports evidence that they represent promising candidates as potential molecular mediators along the causal pathway between smoking to lung function variation. We also detected evidence of colocalization between DNA methylation and lung function at various CpG sites, although not with gene expression. Further work is therefore required to prioritise causal genes at these loci responsible for effects. For example, the functional gene which may be responsible for the association at cg21356710 could be *UBXN2A*, as although it is not the closest gene to the CpG site, it has been previously implicated in nicotine metabolism ^40^. However, future research is necessary to identify strong evidence supporting this.

#### Tissue specificity

A recent study which investigated the colocalization of mQTLs with genetic risk variants for COPD identified several mQTLs in lung tissue which may be involved in COPD pathogenesis ^41^. These findings did not overlap with the findings of this study, perhaps due to differences in tissue type. However, as some of the CpG sites which were causally implicated in our MR analysis may be exerting their effect on lung function via smoking behaviour, this suggests that lung tissue may not always be the most relevant for appraising causal effects. Future work should evaluate and integrate mQTL and eQTL effects from multiple tissues to elucidate causal effects in the most biologically relevant tissues. For example, lung-derived tissue would be ideal to investigate molecular mechanisms which influence lung function as undertaken in our study.

#### Measurement imprecision

One of the main limitations of mediation analysis is the assumption of no measurement error. Mendelian randomization attempts to overcome this limitation with the use of genetic variants which are typically measured with high accuracy. However, differential measurement precision of the phenotypes being investigated in an MR approach can lead to spurious findings in certain instances.

One explanation for the finding that DNA methylation has a causal effect on smoking at several of the CpG sites is that the SNPs used to instrument DNA methylation have their primary effect through smoking. We assessed this using the Steiger test which indicated that this alternative explanation was not likely for those CpG sites where DNA methylation had a causal effect on smoking. However, this test is liable to give inaccurate causal directions if there are large differences in sample size between the two samples, or if the phenotypes have differences in measurement precision ^20^, which is likely to be the case in this context. To assess this further, we compared the magnitude of the mQTL effects in a non-smoking subset of the ARIES cohort (children at age 7) and found similar effects.

#### Strengths

Despite these limitations, this study has several strengths, which include the systematic evaluation of the causal effect of a large number of smoking-related CpG sites on lung function; the replication of findings in different smoking strata and in an independent dataset; the integration of several large-scale datasets to evaluate the causal relationship between smoking, DNA methylation and lung function; the application of a formal two-step MR approach to evaluate mediation; as well as the use of a colocalization approach which integrated gene expression data.

### Conclusions

Using a Mendelian randomization approach, we identified several CpG sites where DNA methylation may have a causal effect on lung function, as assessed by FEV_1_. There was evidence to suggest that the mechanism of action for DNA methylation at some of these sites was via effects on smoking behaviour (rather than vice versa), and also changes in gene expression. Findings highlight potential therapeutic targets for improving lung function and possibly smoking cessation, although further studies with larger-scale and tissue-specific DNA methylation and expression data are required to confirm these results.

## Methods

### mQTL identification: The Accessible Resource for Integrated Epigenomic Studies (ARIES) in the Avon Longitudinal Study of Parents and Children (ALSPAC)

ALSPAC is a large, prospective cohort study based in the south-west of England. A total of 14 541 pregnant women residing in Avon, UK, with expected dates of delivery from 1 April 1991 to 31 December 1992 were recruited and detailed information has been collected on these women and their offspring at regular intervals ^42,43^. The study website contains details of all the data that are available through a fully searchable data dictionary (http://www.bristol.ac.uk/alspac/researchers/our-data/). Written informed consent has been obtained for all ALSPAC participants. Ethical approval for the study was obtained from the ALSPAC Ethics and Law Committee and the Local Research Ethics Committees.

As part of the Accessible Resource for Integrated Epigenomics Studies (ARIES) project ^10,44^, the Illumina Infinium HumanMethylation450 (HM450) BeadChip was used to generate epigenetic data on cord blood and peripheral blood samples from 1,018 mother-offspring pairs in the ALSPAC cohort at five time points (birth, childhood, adolescence, antenatal and middle age). The ARIES participants were previously genotyped as part of the larger ALSPAC study (http://www.bristol.ac.uk/alspac), with quality control, cleaning and imputation performed at the cohort level as described previously ^10^.

Matrix eQTL software ^45^ was used to perform preliminary association analysis of SNPs with CpG sites in the HM450 array with further multivariable linear regression analysis run in PLINK1.07 ^46^ and Genome-wide Complex Trait Analysis (GCTA) performed ^47^ as previously described ^10^. These data are publicly available via an online catalog (http://mqtldb.org) ^10^.

### Genome-wide association of forced expiratory volume and lifetime smoking behaviour: UK Biobank

We used genetic association data from individuals in the UK Biobank. The UK Biobank study is a large population-based cohort of 502,682 individuals aged 37-73 years who were recruited across the UK between 2006 and 2010, with extensive health and lifestyle questionnaire data (including smoking behaviour), physical measures (including spirometry) and DNA samples. The study protocol is available online (http://www.ukbiobank.ac.uk/wp-content/uploads/2011/11/UK-Biobank-Protocol.pdf) and more details have been published elsewhere ^48^. The UK Biobank study was approved by the North West Multi-Centre Research Ethics Committee (reference number 06/MRE08/65) and at recruitment all participants gave informed consent to participate in UK Biobank and be followed-up.

Participants were genotyped either using the Affymetrix UK BiLEVE Axiom array or the Affymetrix UK Biobank Axiom array. Details of how the genotype data were cleaned, imputed and released to the scientific community are detailed elsewhere^49^. Summary-level genetic association statistics for FEV_1_ were obtained from a recent GWAS of FEV_1_ (covariate adjusted and inverse-normal rank transformed) in UK Biobank (n=321,047) ^17^ and for lifetime smoking behaviour, from a GWAS of a comprehensive smoking index metric derived from data on smoking duration, heaviness and cessation in UK Biobank participants (n= 462,690) ^21^.

### Two sample Mendelian randomization: ARIES and UK Biobank

To assess the causal effect of DNA methylation at smoking-related CpG sites on lung function, we conducted two-sample MR ^13^. In this approach, information on the SNP-exposure (here DNA methylation) and SNP-outcome (here lung function (FEV_1_)) effects are derived from genome-wide association analysis conducted in separate studies, using the TwoSampleMR package in R ^50^.

For the smoking-related CpG sites which could be instrumented by mQTLs, we performed a look-up of the identified mQTLs in the lung function GWAS summary data from UK Biobank. We extracted the following summary data for each SNP: the effect estimate for lung function per copy of the effect allele and its standard error, the reference allele, and the effect allele along with its frequency. We combined information on the SNP-lung function associations from UK Biobank with information on the SNP-methylation associations from ARIES in instrumental variable analysis, described below.

For each SNP, we calculated the change in FEV_1_ per standard deviation (SD) increase in methylation by the formula βGD/βGP (also known as a Wald ratio), where βGD is the standard deviation change in volume of air exhaled in one second per copy of the effect allele and βGP is the standard deviation increase in methylation per copy of the effect allele. Standard errors of the Wald ratios were approximated by the delta method ^51^. Where multiple conditionally independent mQTLs were available for the same CpG site, these were combined in a fixed effects meta-analysis after weighting each ratio estimate by the inverse variance of their associations with the outcome (inverse variance weighted (IVW) approach)).

### Replication: SpiroMeta

We attempted to replicate findings regarding the causal effect of DNA methylation using an independent second sample for the two sample MR approach. For this, we used data available on genetic variants and lung function (FEV_1_) in 79,055 individuals of European ancestry from 22 studies, combined in a meta-analysis by the SpiroMeta meta-analysis ^17^.

### Stratification: UK BiLEVE

To investigate the extent to which the genetically-predicted effects of DNA methylation on lung function are modified by smoking status, we conducted an MR analysis stratified by smoking status. For this, GWAS of FEV_1_ has been undertaken in 48,931 individuals from the UK BiLEVE study, a subset of UK Biobank participants who were selected from the extremes of the lung function distribution (extremely low, near-average and extremely high) and by smoking status (never vs heavy smokers (mean of 35 pack years of smoking)) ^52,53^. Genotyping was undertaken using the Affymetrix Axiom UK BiLEVE array for 24,457 smokers and 24,474 non-smokers in UK BiLEVE.

### Sensitivity analysis

#### DNA methylation and lung function: direction of causality

Where there was evidence that DNA methylation may have a causal effect on lung function, we evaluated the possibility of reverse causation, whereby a SNP used to instrument DNA methylation has its primary effect through lung function rather than vice versa. For this, we performed the MR Steiger test ^20^, implemented in the TwoSampleMR package ^50^ using the summary GWAS data from ARIES and UK Biobank previously outlined, to determine the likely direction of effect.

Furthermore, we conducted the reciprocal MR at these CpG sites to appraise the causal effect of lung function (FEV_1_) on DNA methylation. For this, we performed a look-up of 221 SNPs with associations of p<5×10^−8^ from the UK Biobank GWAS of FEV_1_, in GWAS of DNA methylation at the CpG sites of interest in the middle age time point using exact linear regression of methylation beta-values at each CpG site on SNP genotypes, with adjustment for age, sex, top ten ancestry principal components, bisulphite conversion batch and estimated white blood cell counts using PLINK1.07 ^10^.

#### Smoking behaviour and DNA methylation: direction of causality

We also performed bidirectional Mendelian randomization to evaluate the direction of effect between lifetime smoking behaviour and DNA methylation at the identified CpG sites. For lifetime smoking behaviour, we obtained summary statistics for 126 independent SNPs identified in a GWAS of comprehensive smoking index ^21^, with p<5×10^−8^. We performed a look-up of these SNPs in GWAS of DNA methylation at the CpG sites of interest, as described above. We then conducted MR to appraise the causal effect of lifetime smoking behaviour on DNA methylation. We also performed a look-up of mQTLs used to instrument DNA methylation at the CpG sites of interest in the summary data from the GWAS of lifetime smoking behaviour and conducted another two-sample MR analysis to appraise the causal effect of DNA methylation on lifetime smoking behaviour.

#### Negative control

To assess the specificity of the mQTL effect on DNA methylation (not via smoking behaviour), we also assessed the association between the mQTL and DNA methylation at the CpG sites of interest in the childhood time point of ALSPAC using exact linear regression as described above. This can be viewed as a negative control analysis as the association should not be present in this group of non-smoking individuals if it is driven by smoking behaviour.

### Supplementary Analyses

#### Mediation analysis

For those CpG sites where there was consistent evidence that methylation had a causal effect on lung function, and where lifetime smoking was also causally implicated, we first used a two-step MR approach ^12^ to investigate mediation. Prior to this, we performed an MR analysis to estimate the total causal effect of lifetime smoking behaviour on lung function, by performing a look-up of the SNPs associated with lifetime smoking behaviour in the GWAS summary data for FEV_1_.

For those CpGs where there was evidence that smoking influenced DNA methylation which in turn influenced lung function, we used the “product of coefficients” method ^22^ to obtain an estimate for the indirect effect of smoking on lung function via DNA methylation. For those CpGs where there was evidence that DNA methylation influenced smoking which in turn influenced lung function, we used the “product of coefficients” method to obtain an estimate for the indirect effect of DNA methylation on lung function via smoking. This approach is outlined in **Figure 3**. Standard errors for the indirect effect were derived by using the delta method.

Another Mendelian randomization approach which may be used to assess mediation is multivariable MR (MVMR) ^23,24^. This approach can be used to determine the direct effect of an exposure on an outcome, which can be subtracted from the total effect to obtain an estimate for the indirect effect (“difference in coefficients method”) ^22^. We used MVMR to estimate the direct effects of lifetime smoking and the identified CpG sites on lung function by including the genetic instruments for smoking and each CpG site in turn in the multivariable models. Standard errors for the indirect effect were derived using the delta method.

#### Multiple-trait colocalization analysis

For those CpG sites where there was evidence of a causal effect on lung function, we applied multiple-trait colocalization (‘moloc’) ^16^ to investigate whether the variant responsible for influencing methylation at each CpG site was the same variant influencing changes to both nearby gene expression and lung function ^25,26^. We applied moloc using data derived from 3 different sources; mQTL data from the middle age timepoint in ARIES (mean age 47.5), GWAS summary data for FEV_1_ from UK Biobank ^17^ and expression quantitative trait loci (eQTL) data derived from whole blood from the eQTLGen consortium ^54^. We ran moloc multiple times to investigate colocalization with the expression of all genes within 1Mb of the CpG site of interest. Analyses were only undertaken if there were at least 50 variants (MAF ≥ 5%) in common between all 3 datasets. As recommended by the authors of moloc, a posterior probability of association (PPA) of 80% or higher was considered evidence of colocalization. This approach therefore suggests that loci with evidence of genetic colocalization harbour a single causal variant which is responsible for variation in DNA methylation, gene expression and lung function. When there was evidence at the same locus with multiple genes, we reported the association with the highest PPA.

All analyses were undertaken using R (version 3.5.1).

## Data Availability

Please address correspondence and material requests to R.C.R.

## Funding

This work was supported by the Integrative Epidemiology Unit which receives funding from the UK Medical Research Council and the University of Bristol (MC_UU_00011/1 and MC_UU_00011/5). This work was also supported by CRUK (grant number C18281/A19169) and the ESRC (grant number ES/N000498/1). T.G.R. is a UKRI Innovation Research Fellow (MR/S003886/1). R.C.R. is a de Pass Vice Chancellor Research Fellow at the University of Bristol. M.D.T. is supported by a Wellcome Trust Investigator Award (WT202849/Z/16/Z). The research was partially supported by the NIHR Leicester Biomedical Research Centre; the views expressed are those of the author(s) and not necessarily those of the NHS, the NIHR or the Department of Health.

### Acknowledgements

We thank the SpiroMeta Consortium for contributing summary statistics to this work. We would also like to thank Alice Carter, Dipender Gill and Eleanor Sanderson for useful discussions regarding the mediation analysis. This study was made possible with the financial support of Jonathan de Pass and Georgina de Pass.

## Author contributions

R.C.R. and T.G.R. conceived the study; E.J., R.K., T.G.R. and R.C.R. performed the analysis; E.J., T.G.R. and R.C.R. wrote the paper. All authors provided comments on the paper.

## Materials and Correspondence

Please address correspondence and material requests to R.C.R.

## Supplementary figures

**Supplementary figure 1.**
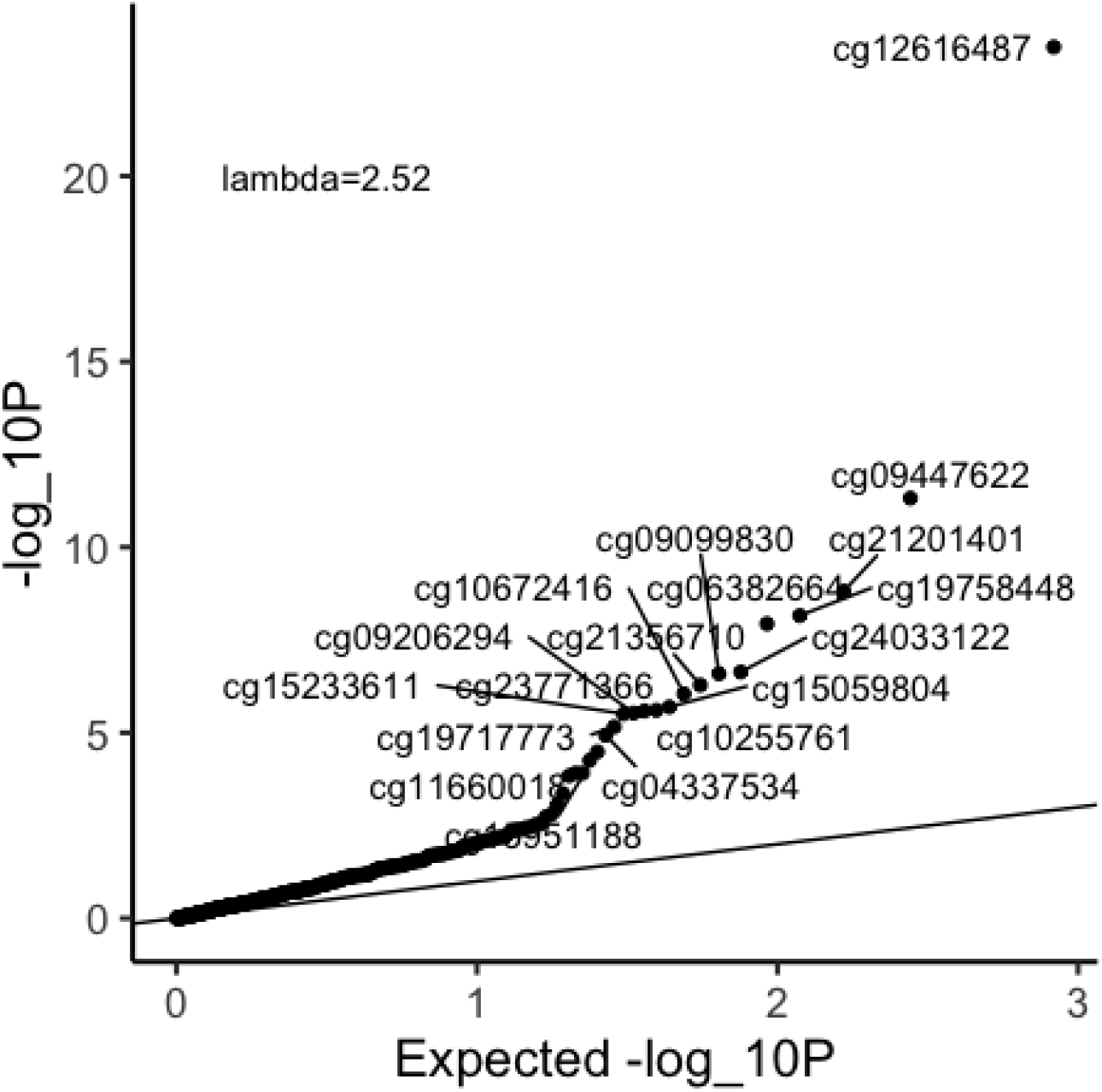
Quantile-quantile plot of the observed vs expected p-values of the associations between smoking-associated DNA methylation and lung function (FEV_1_). The line represents the null hypothesis of no association with lung function. Deviation from the expected distribution of P-values is evident with a lambda of 2.52.

**Supplementary figure 2.**
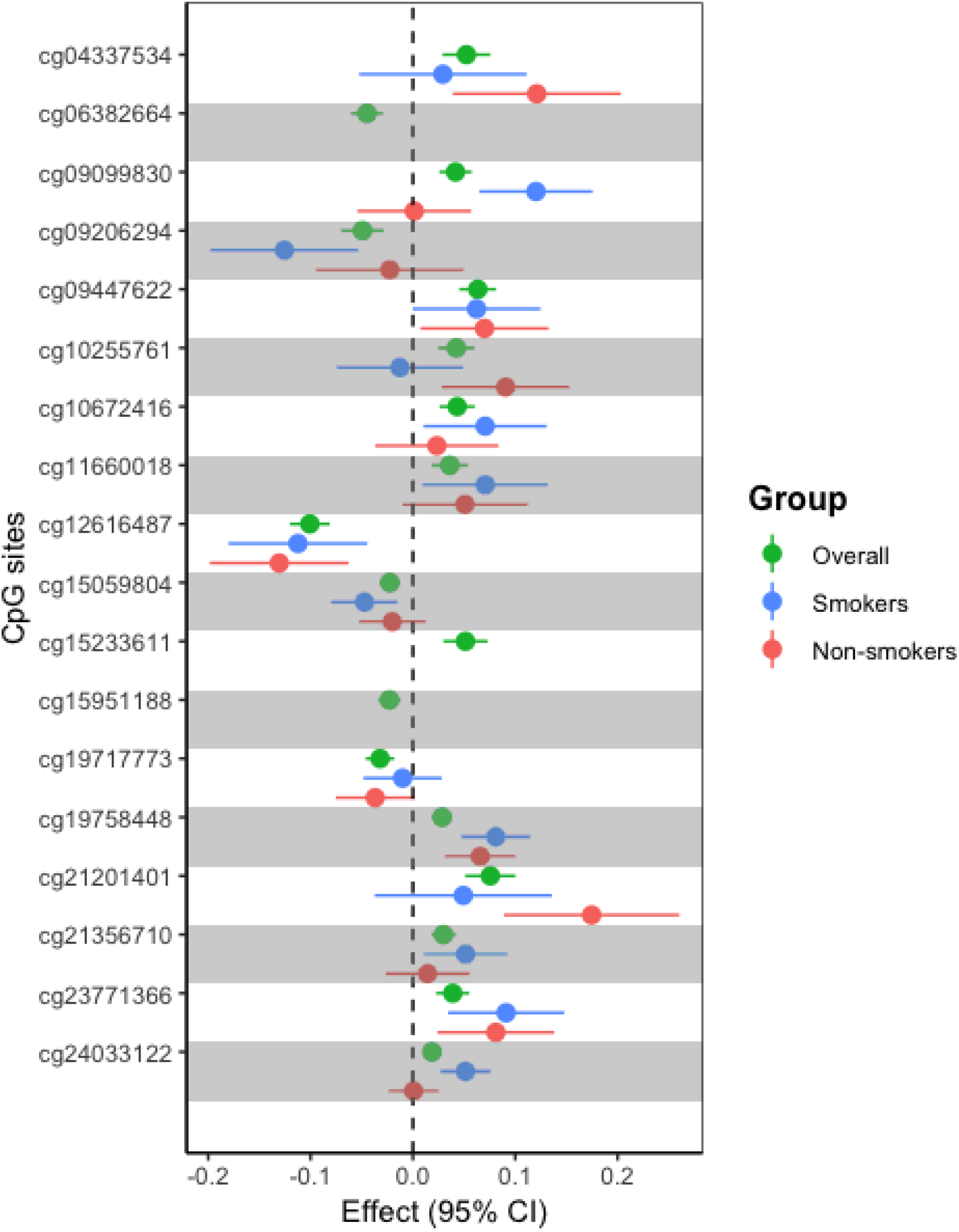
Results of MR analysis of the effect of smoking-associated DNA methylation on lung function (FEV1) stratified by smoking status. Effect sizes and 95% confidence intervals (CI) for each of the top eighteen CpG sites are shown for the overall UK Biobank sample in green, and the smoking and non-smoking UK BiLEVE samples in blue and red, respectively. One CpG (cg15233611) was not available in the UK BiLEVE analysis and so only the result for the UK Biobank analysis is shown.

**Supplementary figure 3.**
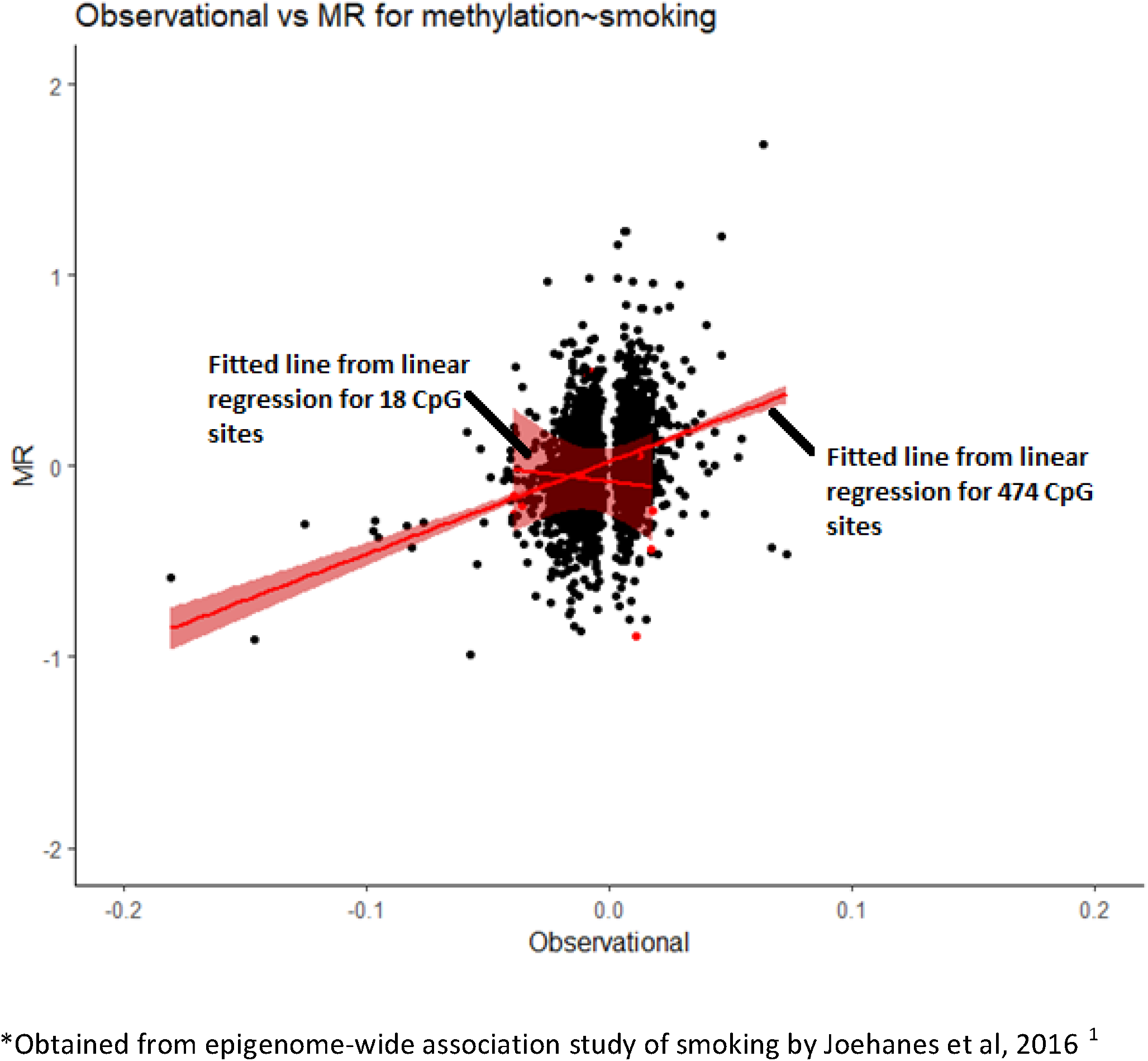
Comparison of observational* and Mendelian randomization effect estimates of smoking on DNA methylation at smoking-related CpG sites

